# Health Outcome Predictive Modelling in Intensive Care Units

**DOI:** 10.1101/2022.12.15.22283527

**Authors:** Chengqian Xian, Camila P. E. de Souza, Felipe F. Rodrigues

## Abstract

The literature in Intensive Care Units (ICUs) data analysis focuses on predictions of length-of-stay (LOS) and mortality based on patient acuity scores such as Acute Physiology and Chronic Health Evaluation (APACHE), Sequential Organ Failure Assessment (SOFA), to name a few. Unlike ICUs in other areas around the world, ICUs in Ontario, Canada, collect two primary intensive care scoring scales, a therapeutic acuity score called the “Multiple Organs Dysfunctional Score” (MODS) and a nursing workload score called the “Nine Equivalents Nursing Manpower Use Score” (NEMS). The dataset analyzed in this study contains patients’ NEMS and MODS scores measured upon patient admission into the ICU and other characteristics commonly found in the literature. Data were collected between January 1st, 2015 and May 31st, 2021, at two teaching hospital ICUs in Ontario, Canada. In this work, we developed logistic regression, random forests (RF) and neural networks (NN) models for mortality (discharged or deceased) and LOS (short or long stay) predictions. Considering the effect of mortality outcome on LOS, we also combined mortality and LOS to create a new categorical health outcome called LMClass (short stay & discharged, short stay & deceased, or long stay without specifying mortality outcomes), and then applied multinomial regression, RF and NN for its prediction. Among the models evaluated, logistic regression for mortality prediction results in the highest area under the curve (AUC) of 0.795 and also for LMClass prediction the highest accuracy of 0.630. In contrast, in LOS prediction, RF outperforms the other methods with the highest AUC of 0.689. This study also demonstrates that MODS and NEMS, as well as their components measured upon patient arrival, significantly contribute to health outcome prediction in ICUs.

## 1 Introduction

The Intensive Care Unit (ICU) is a unique hospital department, providing the highest level of treatment for a hospital’s highest acuity patients. It is an intrinsically technological environment where each patient generates thousands of data points per day, and data-driven management applied to ICU allows not only an evaluation of ICU performance but has other implications, including better planning of scarce resources and transition of care and discharge [1, 2]. ICU scoring system is an essential tool to describe the severity of patients’ illnesses to improve clinical decision-making and predict patients’ health outcomes [3–7]. Existing scoring systems to assess patient illness severity on admission to ICU include the Acute Physiology and Chronic Health Evaluation II and its variations (APACHE II) [8] as well as the Simplified Acute Physiology Score (SAPS II) [9]. In addition to describing the severity of a patient’s disease, Multiple Organ Dysfunction Score (MODS) [10] and Sequential Organ Failure Assessment Score (SOFA) [11] were also developed to specifically evaluate patients’ organ function or determine the rate of organ failure. The MODS score (see the full list of its components in Appendix Table A6) is scaled from 0 to 24, and it is constructed from six organ systems and demonstrates a strong correlation with the risk of ICU mortality [10]. The Nine Equivalents Nursing Manpower Use Score (NEMS) [12] (see the full list of its components in Appendix Table A7) was developed from the Therapeutic Intervention Scoring System (TISS) [13] to measure the nursing workload in ICU. NEMS is based on nine life support interventions ranging from 0 to 56, and has been validated in an adult 30-bed medical-surgical ICU in a tertiary care university hospital. Its good agreement is further confirmed with TISS-28 [14].

In Ontario, Canada, hospitals collect MODS and NEMS for reporting purposes, but they lack the necessary measurements to calculate widely used severity scores like APACHE and SAPS [15]. Limited recent studies have explored the relationship between MODS, NEMS, and the health outcomes of ICU patients in Ontario. Notably, two main studies conducted by Kao et al. [16] and Rodrigues [15] provide valuable insights. In their study, Kao et al. developed a logistic regression model to predict patient mortality using MODS, NEMS, and general patient characteristics, such as age, based on a data set comprising 8, 822 patients collected from Ontario between January 1, 2009, and November 30, 2012. Rodrigues expanded on Kao et al.’s work in two ways using more recent data collected between January 1, 2015, and December 31, 2016, which included 4, 758 patients. First, Rodrigues enhanced the mortality prediction models by incorporating the components of MODS and NEMS, not only through logistic regression but also utilizing supervised machine learning methods like random forests and neural networks. Additionally, Rodrigues focused on developing models for predicting length of stay (LOS) in the ICU, as it significantly influences ICU resource planning due to the strong correlation between ICU costs and LOS [17]. The key distinction between these two studies lies in their research focus. Rodrigues primarily compares the performance of multiple statistical and supervised machine learning algorithms in predicting mortality and LOS, while Kao et al. primarily investigate statistically significant predictors in distinguishing patients at low and high risk of mortality.

In this work, we expand the work of [16] by adding the components of MODS and NEMS as predictors. We then consider logistic regression and compare it with two other common machine learning methods (random forests and neural networks) for mortality or LOS predictions. We validate the work of [15] using a larger data set with 15, 350 patients collected from the same ICUs between Jan. 1, 2015, and May 31, 2021. Our main research objectives are: (i) investigate the significance of MODS and NEMS along with their components in both mortality and LOS predictions; (ii) analyze the quantitative effect of patient general characteristics and the admission characteristics (e.g., MODS and NEMS) via regression models; (iii) construct a new categorical outcome, LMClass (LOS-Mortality Class), to combine mortality and LOS, and fit multinomial regression models, random forests, neural networks for its prediction.

As we will demonstrate and discuss, our proposed models with MODS and NEMS components added as predictors significantly improve the performance of mortality and LOS predictions. Compared with [15], our second objective fills the gap in the interpretation of risk factors on health outcomes predictive modelling and provides a better understanding of a predictive model. More sophisticated methods like machine learning algorithms may perform better in classifying health outcomes. However, their complex model structures make it harder to understand and may lose interpretation power [18]. Additionally, the motivation for analyzing LMClass comes from the “endogeneity” of mortality in ICU length of stay prediction [15, 19]. For instance, deceased patients may have shorter or longer LOS than their discharged counterparts. Ignoring this endogenous effect may cause bias in LOS prediction [20]. Combining these two outcomes may provide a more comprehensive and useful assessment of patient outcomes [21]. We, therefore, suggest a new categorical health outcome, LMClass, with three categories: short stay & discharged; short stay & deceased; and long stay without specifying mortality outcomes. We will discuss the definition of a prolonged LOS in Section 3.1. In this composite outcome, we focus on mortality outcomes in patients with short LOS for short-term allocation and planning of ICU resources.

The rest of the paper is structured as follows. We provide a general literature review on health outcome predictive modelling in ICU in Section 2. Section 3 presents the material and the statistical methodology. Then, Section 4 describes our analysis results. Finally, the conclusion and discussion are presented in Section 5.

## 2 Literature review

With advances in information technology and data science, statistical models and machine learning methods have been applied to ICU data for mortality and LOS predictions [1, 22–25]. In what follows, we present a comprehensive literature review of mortality prediction in Section 2.1 and LOS prediction in Section 2.2.

### 2.1 Mortality prediction

ICU mortality prediction, involving the classification of patients as either discharged or deceased, is commonly approached as a binary classification problem. Extensive research has been conducted for ICU mortality prediction. For comprehensive insights in this field, systematic reviews by Fusaro et al. [26] and Keuning et al. [27] are valuable resources.

Logistic regression combined with the likelihood ratio test (LRT) is widely used to predict patient mortality in ICU [16, 28–36]. In a recent study [37], logistic regression was employed to detect risk factors associated with ICU survival during the COVID-19 pandemic. As one of the most commonly used generalized linear models, logistic regression has an excellent interpretation power via odds ratio [38, 39], which helps quantitatively describe the impact of each predictor in mortality risk [18].

Machine learning algorithms are widely acknowledged as alternatives to logistic regression in various domains, including ICU mortality prediction. These algorithms encompass a range of methods, such as decision trees, support vector machines, k-nearest neighbors, random forests, super learners, boosting, and neural networks, among others [15, 40–44]. Furthermore, there exists an extensive body of literature that explores the application of machine learning methods to predict ICU mortality outcome specifically for COVID19 patients [45–49]. When reviewing these studies, it is essential to consider several unique characteristics of the analyzed ICU data. These factors include the country or region from which the data originates, the patient population under study (e.g., adults or children, patients with severe pneumonia or heart disease), and whether the data is sourced from a single center or multiple centers. By taking these factors into account, researchers can better contextualize and interpret the findings from these diverse studies.

### 2.2 LOS prediction

When reviewing the literature on ICU length-of-stay (LOS) prediction, we can categorize the existing studies into three main groups: typical regression analysis, binary classification, and survival regression. In typical regression analysis, two common approaches are employed: multiple linear regression (MLR) and regression using machine learning techniques. For binary classification, the first step is to define what constitutes a prolonged LOS. Researchers need to establish a threshold or criteria for defining a long stay in the ICU. Once this definition is established, classification methods are applied to predict whether a patient will have a prolonged LOS based on the available features and predictors. Survival regression analysis is another approach used in LOS prediction, specifically designed to handle censoring in the data collection process. It is worth noting that different scoring systems may be employed in various studies, depending on the geographical location of the ICUs and the specific context of the research. Recent comprehensive reviews on LOS prediction can be also found in [50–52].

Zimmerman [7] developed an MLR procedure using APACHE IV to estimate ICU stay using data across ICUs in the United States. They showed that the accuracy and utility of the predictions based on the APACHE IV model were unsatisfactory. In [53], researchers improved patient LOS prediction by firstly optimizing a threshold for a prolonged stay and building a multivariate linear regression with the severity score information on day five, achieving a better prognosis than that based on ICU day one information alone. Similar studies where the MLR was applied can be found in [28, 54, 55]. Regression models based on machine learning are also widely considered and built, which include support vector regression [56], gradient boosting regression [57], random forests regression [58, 59].

Sometimes, clinical practitioners are also interested in the binary classification for LOS prediction (long-stay or short-stay), and therefore classification methods including logistic regression, support vector machine, random forests, and neural networks were also implemented in predicting prolonged LOS or short LOS [15, 56, 60, 61]. Neural networks were developed as predictive instruments for ICU LOS for the first time in [60]. Defining prolonged LOS as a stay greater than two days, they found that the neural networks model performed well with an area under the receiving operating characteristic curve (AUC) of 0.6960 in the validation set. In [56] and [15], the authors performed similar work on LOS prediction by applying machine learning methods but based on different severity scores (SOFA score in [56] while MODS in [15]).

Recently, survival regression analysis is also conducted for LOS prediction, where the time of ICU stay is considered a survival time response to correct for censoring. The AFT model with Weibull distribution was developed in [15] for short-term capacity planning in ICUs from Ontario, Canada. The Weibull AFT model was also applied in [62, 63] to investigate the effect of predictors on LOS for COVID-19 patients in the UK. Authors in [62] further built the log-normal AFT compared to the one with Weibull distributional assumption. The Cox PH model was developed in [64] to analyze the effect of severity scores, SOFA, on LOS for COVID-19 patients in India.

## 3 Material and methods

### 3.1 Data source and data management

Our research is a retrospective study conducted at two teaching hospital ICUs in southwestern Ontario, which specialize in the care of various patient populations, including neurosurgical, cardiovascular surgery, and transplantation patients. Data were collected from Jan. 1, 2015, to May 31, 2021 and stored in four separate data sets called MODS, NEMS, Source and Awaiting Transfer. MODS is the data set containing the MODS score along with its components measured upon patient admission to ICU. NEMS is another important set containing patients’ NEMS scores and their components measured daily in ICU. ICU discharge time and destination are also provided in MODS and NEMS data sets. The Source set includes de-identified patient general characteristics (e.g., age, sex) and admission characteristics (e.g., admission source, admission diagnosis, patient category, referring service). The last data set, Awaiting Transfer, provides the admission time and the awaiting transfer discharge start date time, both of which were used to calculate the clinical LOS.

Since we have four separate data sets, several new variables were created in each set before merging them into a single data set. In the MODS set, we created *Mortality* as a binary response which can be constructed from discharge destination: 1 if the patient is deceased at the end of the ICU stay, otherwise 0 for being discharged alive. Besides, we calculated the *total LOS*, defined as the period between patients’ admission to and exit from ICU, which is used to detect extreme values of stay in data cleaning procedure as done in [15]. In the Source set, we edited the admission source and admission diagnosis by combining some of their categories in the same way proposed in [15]. For admission sources, we kept the Emergency Department, Operating Room, and Unit/Ward/Stepdown while combining the other levels to Outside Hospital/Other. For admission diagnosis, we kept the Cardiovascular/Cardiac/Vascular, Gastrointestinal, Neurological, Respiratory, and Trauma while combining other levels to Other. In the Awaiting Transfer set, we calculated the *clinical LOS*, defined as the period between patient admission to ICU and the physician’s disposition decision (i.e., transfer or discharge). Then the prolonged LOS called *IsLong* is defined as a stay longer than five days, based on the empirical distribution of LOS as discussed in Section 4.1. In other words, *IsLong* takes the value of 1 if clinical LOS *>* 5 days and 0 otherwise. Besides, *LMClass*, a categorical response with three levels, was also created by combining *Mortality* and *IsLong* : short stay & discharged, short stay & deceased, or long stay without specifying mortality outcomes.

To build predictive models for each health outcome, we need to combine these four separate data sets into a single one. Patients’ ID and admission time can link these four data sets. We first extracted the MODS score with its components from the MODS set and used patients’ IDs and admission time to merge the NEMS score with its components on admission day in the NEMS set. To obtain the clinical LOS, patient characteristics, and admission characteristics, we merged the latest combined data set with the Awaiting Transfer set and the Source set, resulting in a single data set with 15,474 cases. Similar to [15], some cases with large total LOS (*≥* 60 *days*, 90 cases), unknown or missing sex (15 cases), and unusual age (*≥* 110 *years*, 19 cases) were removed from our merged data set for further analysis, resulting in a finalized data set with 15,350 cases. A flow chart of this process is provided in Figure 1.

**Fig. 1:**
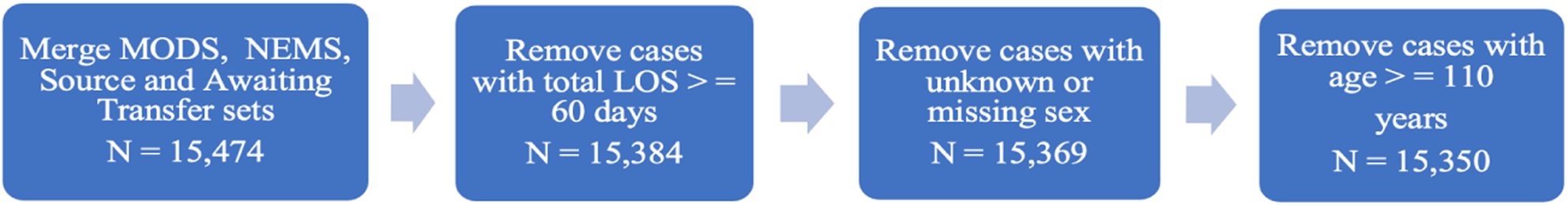
Flow chart of data cleaning

### 3.2 Statistical analysis

The finalized data set (*N* = 15, 350) was split into a training set (*N* = 10, 745) and a validation set (*N* = 4, 605) with a ratio of 7:3. Each proposed model was built on the training set and validated on the validation set. Figure 2 presents a flow chart of ICU health outcome predictive modelling. To present more details of our methodology and assure reproduciblility, we fill in the scorecards suggested by [65] for mortality or LOS prediction (Table A1 in Appendix A) and LMClass prediction (Table A2).

**Fig. 2:**
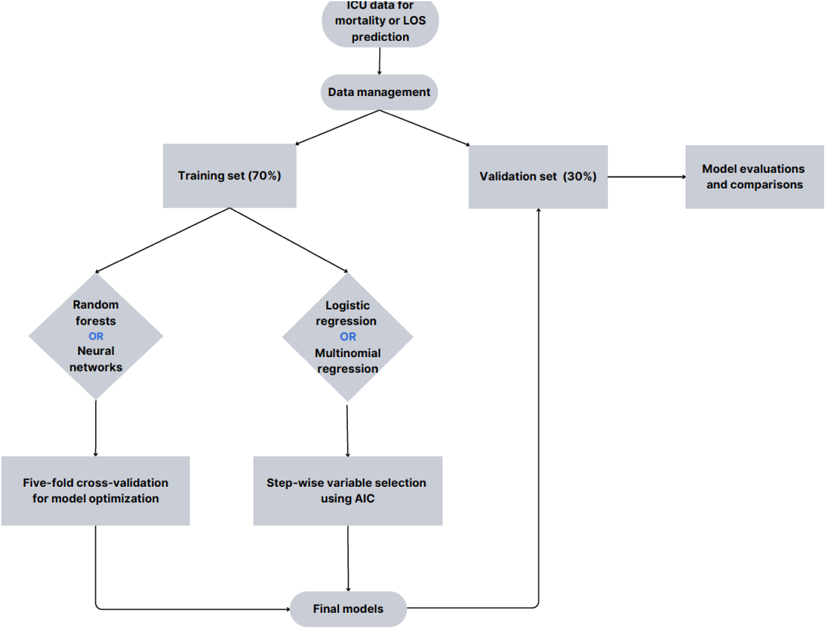
Flow chart of ICU health outcomes predictive modelling

Logistic and multinomial regression are two widely used generalized linear models for modelling binary and multi-class responses, respectively [18]. As we illustrated in our objectives, we aim to apply the odds ratio with respect to a unit change from the risk factor to describe the corresponding effect on the health outcome. For example, in mortality prediction, considering a fitted logistic regression model on a series of risk factors including MODS with a positive estimated regression coefficient denoted by *β*^^^, we can describe as follows: with other risk factors unchanged, an ICU patient with one unit increase in MODS score will increase the odds of death by 100 exp *β*^^^ *−* 1 % [18].

Random forests (RF), proposed in [66], is another popular model for classification by constructing a multitude of decision trees. The two most critical parameters in the random forests model are the number of trees to be built and the number of variables randomly sampled as candidates at each split. In the R package *RandomForest* [67], these parameters are represented by function arguments *ntree* and *mtry*, respectively. In our data set, there are 23 predictors, so *mtry* can be 1, 2*, …,* or 23. For the number of trees, we chose from 100, 500 and 1000. Combining both *ntree* and *mtry*, we have 69 (i.e., 23 *×* 3) alternative models built on the training set using all predictors available. To analyze the black-box mechanisms of random forests, one of the most efficient variable importance measures, mean decrease accuracy (MDA) introduced in [68], can be applied, which is a method of computing the predictor importance on permuted out-of-bag samples based on the mean decrease in the accuracy. In other words, if MDA is high for a predictor, this predictor is important. Visualization of MDA for all predictors is provided after fitting an RF model in the same R package, *RandomForest*.

Neural networks (NN) were built based on the resilient back-propagation with weight backtracking algorithm proposed by Riedmiller M. in 1994 [69]. Before modelling, we conducted data preprocessing for numeric predictors (e.g., age) and ordinal categorical predictors (e.g., components of MODS) by min-max normalization and for nominal categorical predictors (e.g., admission diagnosis) by a one-hot encoding scheme [70]. The two most important parameters of the NN are the number of hidden layers and the number of neurons on each layer. We consider one or two hidden layers with one to five neurons, and as a result, we need to find the optimal NN model from 30 (i.e., 5 + (5 *×* 5)) alternative combinations of different numbers of hidden layers and neurons. All statistical analyses were performed using software R version 4.2.1.

In what follows, the evaluation metrics for the classification are demonstrated. In mortality and LOS binary predictions, we assessed model discrimination performance by AUC from the receiver operating characteristic (ROC) curve, sensitivity (Sen, also called “recall”), specificity (Spe), accuracy (Acc), Matthews correlation coefficient (MCC), positive predictive value (PPV, also called “precision”), negative predictive value (NPV) and F1 score. In LMClass prediction, which is a three-class classification problem, we calculate the accuracy (i.e., the overall percentage of cases correctly classified), the balanced accuracy (i.e., the average of recalls from each class) and the Kappa statistic [71] to evaluate the performance of each proposed model. In binary classification, we use the AUC as a criterion for parameter optimization in RF and NN models, while we use the Kappa statistic for LMClass classification. As discussed in [72], AUC evaluates the overall diagnostic performance of a binary classification and helps select the optimal threshold for determining the presence or absence of a specific health outcome. Furthermore, the Kappa statistic in multi-class classification is an appropriate metric to account for class imbalance [73].

## 4 Results

In this section, we first present the descriptive analysis results of our data set, and then the results regarding mortality, LOS, and LMClass prediction, respectively. We also quantitatively elaborate on how MODS and NEMS affect the prediction of health outcomes via the odds ratio (i.e., the relative risk ratio) from regression-based models.

### 4.1 Descriptive analysis

In our dataset, 11, 963 admitted patients were discharged alive when exiting the ICU while 3, 387 patients died. Most patients stayed less than 5 days in ICU, accounting for 70% of the study population. Table 1 shows the general characteristics of patients (excluding the components of MODS and NEMS) in the training and validation sets. The median with interquartile interval (IQI) was presented for numeric variables, and for categorical variables, raw counts and percentages were presented. We can observe that the median values of MODS and NEMS scores are the same in the training and validation sets (5 points with IQI 3-7 and 32 points with IQI 27-39, respectively). The mortality rate in training is 21.83%, 0.78% lower than that in the validation set. Median clinical LOS in training and validation is 2.545 days (IQI 1.082-5.776) and 2.537 days (IQI 1.115-5.878), respectively.

**Table 1:** Characteristics of ICU admitted patients in the training and validation sets.

| Variables | Training | Validation |
| --- | --- | --- |
| MODS Score (median [interquartile interval] <sup>1</sup>) | 5 [3, 7] | 5 [3, 7] |
| NEMS Score (median [interquartile interval]) | 32 [27, 39] | 32 [27, 39] |
| Mortality, yes, (N, [%] <sup>2</sup>) | 2346 [21.83%] | 1041 [22.61%] |
| Total LOS, days, (median, [interquartile interval]) | 3.361 [1.671, 6.913] | 3.372 [1.645, 6.885] |
| Clinical LOS, days, (median, [interquartile interval]) | 2.545 [1.082, 5.776] | 2.537 [1.115, 5.878] |
| Age, years, (median, [interquartile interval]) | 63.39 [51.14, 73.46] | 63.55 [50.94, 73.78] |
| Sex, Male, (N, [%]) | 6269 [58.34%] | 2690 [58.41%] |
| Patient Category |  |  |
| Medical, (N, [%]) | 6769 [63.00%] | 2855 [62.00%] |
| Surgical, (N, [%]) | 3976 [37.00%] | 1750 [38.00%] |
| Admission Source |  |  |
| Emergency Department, (N, [%]) | 3740 [34.81%] | 1551 [33.68%] |
| Operating Room, (N, [%]) | 2045 [19.03%] | 922 [20.02%] |
| Unit/Ward/Stepdown, (N, [%]) | 2214 [20.60%] | 960 [20.85%] |
| Outside Hospital/Other, (N, [%]) | 2746 [25.56%] | 1172 [25.45%] |
| Admission Diagnosis |  |  |
| Cardiovascular/Cardiac/Vascular, (N, [%]) | 1754 [16.32%] | 750 [16.29%] |
| Gastrointestinal, (N, [%]) | 988 [9.19%] | 417 [9.06%] |
| Neurological, (N, [%]) | 1473 [13.71%] | 561 [12.18%] |
| Respiratory, (N, [%]) | 3111 [28.95%] | 1336 [29.01%] |
| Trauma, (N, [%]) | 832 [7.74%] | 370 [8.03%] |
| Other, (N, [%]) | 2587 [24.08%] | 1171 [25.43%] |

<sup>1</sup>for numeric variables, the median with interquartile interval was presented;
<sup>2</sup>for categorical variables, we presented raw count (N) and the percentage (%) of each category indicated by an indentation.

Figure 3 shows the histogram with the estimated density function (the red curve) of clinical LOS. There are only a few cases with clinical LOS longer than 20 days and most of cases have clinical LOS between 0 and 4 days, resulting in a right-skewed distribution. Specifically, 30% of ICU patients in the data set stayed longer than 5 days and 15% longer than 10 days. Teaching hospitals in Ontario define a prolonged ICU LOS as longer than 21 days [15], which accounts for 4.1% in our data set. Therefore, we need to adjust the definition of a prolonged LOS to preserve clinical significance while avoiding an extremely imbalanced data set. We consider a stay longer than 5 days (i.e., the 70th percentile) as a prolonged stay in our data set. First, patients may require 2day stay in ICU for routine postoperative monitoring [74, 75]. Second, most standard classification machine learning methods including random forests face great challenge in presence of imbalanced data [76]. It is important to note that different studies may define prolonged LOS using different thresholds, for example, 3-day [77], 5-day [78, 79], 7-day [80], 8-day [81], and 21-day [82].

**Fig. 3:**
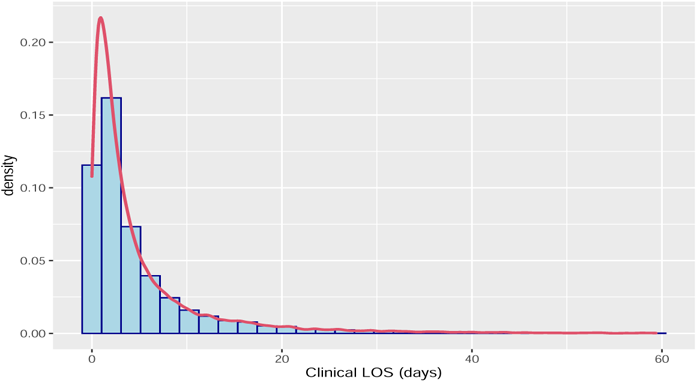
Histogram plot with estimated density (red curve) of clinical LOS.

### 4.2 Mortality prediction

The best RF model in mortality prediction is with *mtry* = 3 and *ntree* = 1000. In other words, the number of variables randomly sampled as candidates at each split on the tree is three, and 1000 trees were built to construct the forests. The best NN model has one hidden layer on which there are two neurons.

Performance of logistic regression, RF and NN on the validation set for mortality prediction is shown in Table 2. ROC curves for all models in the validation set are also presented in Figure 4 which shows no big difference among different models. Logistic regression outperforms RF and NN with the highest scores in AUC (0.795), accuracy (0.705), F1 score (0.532) and PPV (0.415). RF performs the best in achieving the highest sensitivity of 0.748 and NPV of 0.904. Logistic regression has the second highest sensitivity (0.743), while NN provides a relatively lower sensitivity (0.732). The MCC values are low, ranging from 0.364 to 0.373 and the F1 scores are better, ranging from 0.527 to 0.532.

**Fig. 4:**
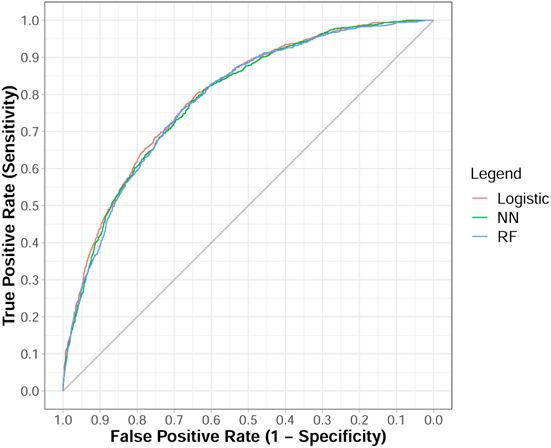
ROC curves for mortality prediction in the validation set. Red: logistic regression; Green: random forests; Blue: neural networks

**Table 2:** Performance of logistic regression, RF and NN on the validation set for mortality prediction.

| Model | AUC | 95% CI | Cutoff | Sen | Spe | Acc | MCC | F1 | PPV | NPV |
| --- | --- | --- | --- | --- | --- | --- | --- | --- | --- | --- |
| Logistic | 0.795 | (0.780, 0.810) | 0.206 | 0.743 | 0.694 | 0.705 | 0.372 | 0.532 | 0.415 | 0.902 |
| RF | 0.788 | (0.773, 0.803) | 0.186 | 0.748 | 0.690 | 0.703 | 0.373 | 0.532 | 0.413 | 0.904 |
| NN | 0.789 | (0.774, 0.804) | 0.221 | 0.732 | 0.695 | 0.703 | 0.364 | 0.527 | 0.412 | 0.899 |

AUC: area under the curve; 95% CI: 95% confidence interval for AUC; Cutoff: optimal threshold for determining mortality obtained via Youden's J statistic [83]; Sen: sensitivity; Spe: specificity; Acc: accuracy; MCC: Matthews correlation coefficient; F1: F1 score; PPV: positive predictive value; NPV: negative predictive value.

In mortality prediction, the F1 scores from the proposed models are slightly larger than 0.5, which, one may think, is close to a random guess. The F1 score in binary classification takes into account both PPV (i.e., precision) and sensitivity (i.e., recall) to provide a more robust evaluation in case of data imbalance. In our predictive modelling for patient mortality, we obtained a lower PPV, around 0.4, while a higher sensitivity, around 0.7, from each proposed model. Therefore, the F1 score is around 0.5. In principle, an F1 score of 0.5 indicates a lower prediction performance. However, such an F1 score resulting from a higher sensitivity still carries meaning for this type of application. For example, since we define death as the outcome of interest, a higher sensitivity means the model correctly detects most patients who will actually die. Moreover, if our model predicts a patient’s death, the patient is still likely to survive based on our PPV values. In contrast, due to our high NPV values, a survival prediction is likely a true negative, i.e., the patient survives.

Table A3 in the Appendix A shows the results of selected predictors for logistic regression based on AIC. Those selected predictors include six components of MODS (Haematologic, Hepatic, Renal, Cardiovascular, Neurologic and Respiratory), NEMS score, six components of NEMS (Basic Monitoring, Intracranial Pressure Monitor, Dialysis, Intra-Aortic Balloon Pump, Other Interventions Within this Unit and Interventions Outside this Unit), Age, Sex, Patient Category, Admission Source and Admission Diagnosis. The odds ratio and its 95% confidence interval (CI) were also calculated for each selected predictor. We find that MODS score is not selected in the optimal model but its six components are. However, NEMS is selected in the model and has an odds ratio of 1.07 (95% CI = [1.06, 1.08]), which indicates that one point increase in NEMS score would increase the relative risk of mortality (i.e., death) by 7% (95% CI = [6%, 8%]), holding the other covariates fixed.

The significant variables returned by the regression model and the important variables from random forests have a high degree of agreement. In logistic regression, the likelihood ratio test (LRT) was applied to all the selected predictors to assess their significance [18], and the corresponding *p*-values were reported in the A3. Except for one of the NEMS components, the Intra-Aortic Balloon Pump, we find that all other selected predictors are statistically significant for mortality prediction. We present a visualization of predictor importance using MDA from the RF model in the left plot of 5. The plot shows that NEMS, MODS, ICU admission source, age, neurological level, patient category and ICU admission diagnosis are the seven most important predictors with an MDA higher than 30%. In addition, these seven predictors are all significant predictors in the logistic regression model for mortality prediction. Practitioners may be interested in the sensitivity (i.e., recall) of the fitted model for mortality prediction. Specifically, they are concerned about the proportion of correct prediction for those patients who deceased at the end of ICU stay. In mortality prediction, the sensitivity coming from RF is 0.748, meaning that it works relatively well in predicting the mortality outcome in those patients who deceased in the end and 74.8% could be correctly predicted. NPV is also an important index in mortality prediction. RF has the highest NPV of 0.904, meaning that if we predict that someone will be discharged alive at the end of the stay, they would likely be discharged with a probability of 90.4%.

### 4.3 LOS prediction

In LOS prediction, the optimal RF model has the same parameter structure as that in mortality prediction where three predictors were randomly sampled at each split on the tree and 1000 trees were built. The best NN model has one hidden layer with one neuron. This simplest NN model was also presented and investigated as the best model in [84] for survival prediction in the ICUs, which is consistent with the empirical analysis result that reducing the complexity of a neural network structure may provide a better performance of prediction for health outcomes in ICUs [85, 86].

Table 3 presents the evaluation measures of all models for LOS prediction in the validation set. The AUC values of all models are lower than those in mortality prediction by around 10%. The MCC values ranging from 0.247 to 0.251 are very low while the F1 scores ranging from 0.503 to 0.508 are still acceptable. On the whole, logistic regression and RF outperform NN. Specifically, logistic regression has the highest score in sensitivity (0.673), MCC (0.251) and NPV (0.820), while RF has the highest score in AUC (0.689), specificity (0.622), accuracy (0.630) and PPV (0.411). The corresponding ROC curves were shown in Figure B1 in Appendix B and once again, no big difference among models can be seen from the plot.

**Table 3:**
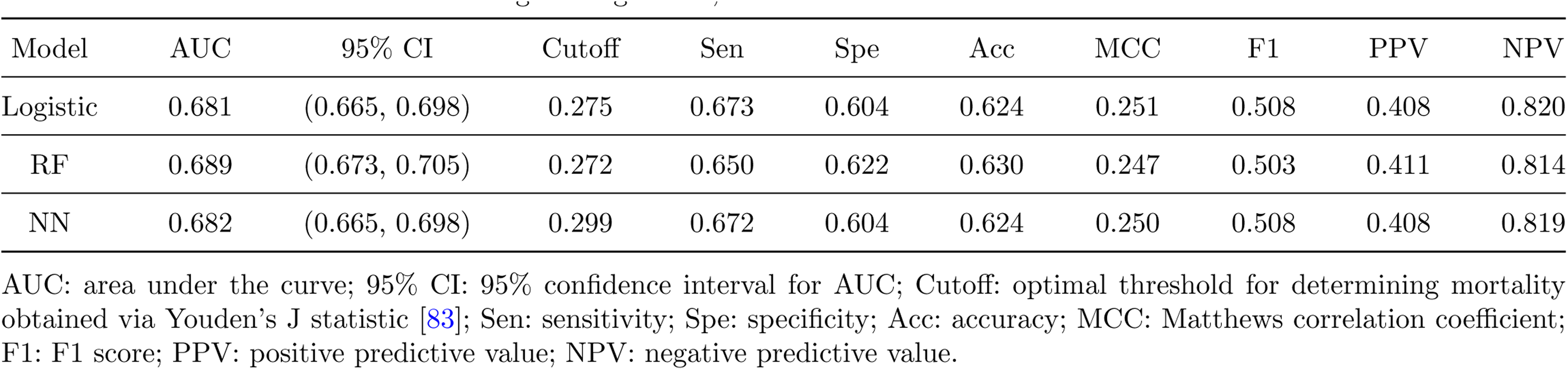
Performance of logistic regression, RF and NN on the validation set for LOS Prediction.

In LOS prediction, sensitivity is one of the most important indices for the choice of models. Logistic regression has the highest sensitivity of 0.673, indicating that 67.3% of patients who stayed more than 5 days could be correctly predicted. The negative predictive values (NPV) from logistic regression is also the highest (0.82), meaning that if we predict one stays less than 5 days, they would probably stay less than 5 days with a probability of 0.82.

Table A4 in Appendix A contains information of the selected predictors for logistic regression in LOS prediction. MODS score with its two components (Cardiovascular and Respiratory), NEMS score with its five components (Central Venous Line, Arterial Line, Intracranial Pressure Monitor, Dialysis and Interventions Outside this Unit), Sex, Admission Source and Admission Diagnosis are selected in the best model. The odds ratios for MODS and NEMS are 1.04 (95% CI = [1.02, 1.06]) and 1.04 (95% CI = [1.04, 1.05]), respectively. This indicates that one point increase in MODS score or in NEMS score would increase the relative risk of staying more than 5 days by 4%, holding the other predictors fixed. Furthermore, from the right plot of Figure 5, we can see NEMS and MODS are both important predictors in prolonged LOS prediction, which is consistent with results of LRT for testing the significance of MODS and NEMS based on logistic regression.

**Fig. 5:**
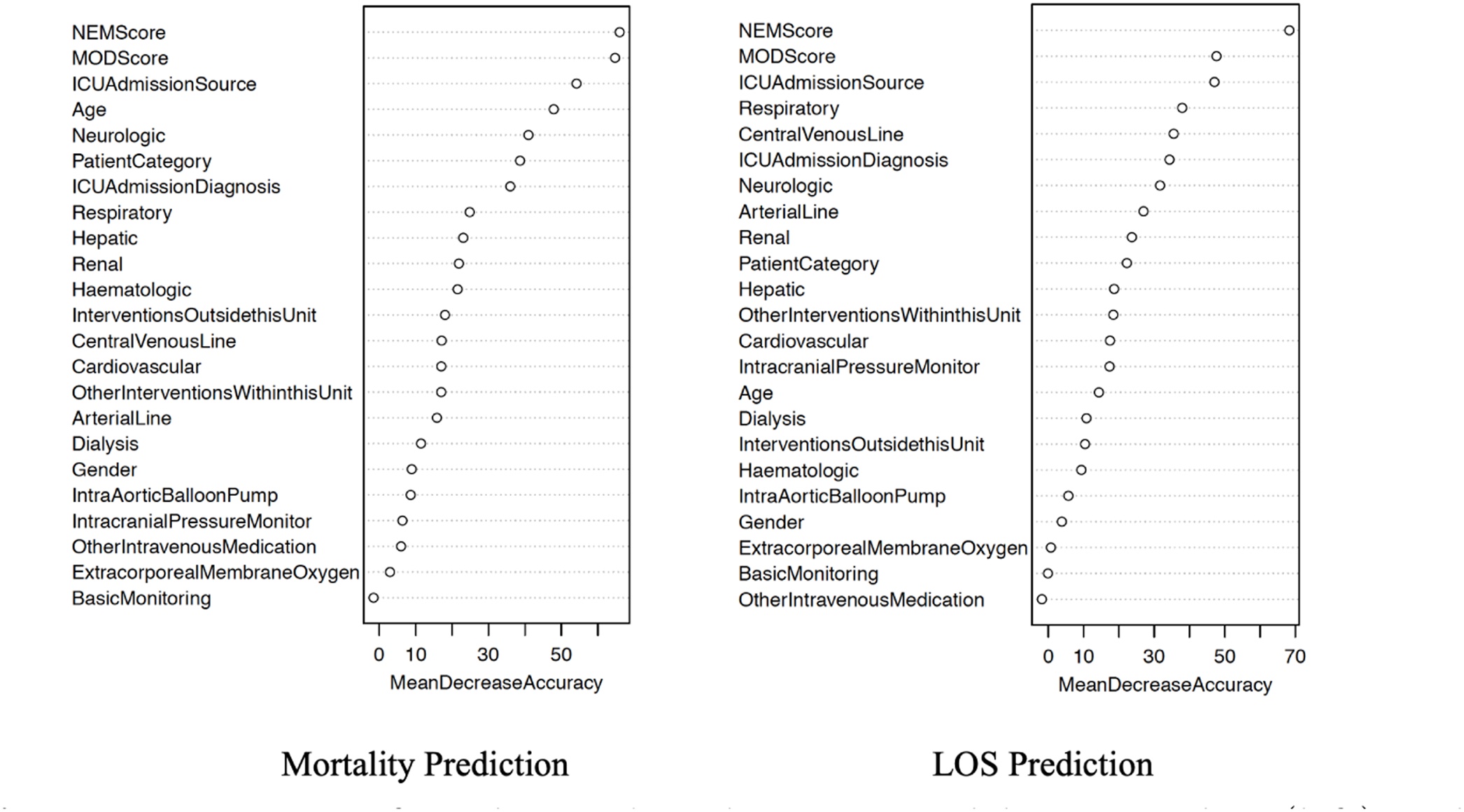
Importance of predictors based on RF model in mortality (left) and LOS (right) predictions.

### 4.4 LMClass prediction

In LMClass prediction, only NN models with one hidden layer reliably converged. Therefore, we considered one to ten neurons within the hidden layer and found that three neurons yielded the highest Kappa statistic. For the RF model fitting, the optimal number of random predictors at each split and the optimal number of trees built are 9 and 500, respectively.

Table 4 presents each overall accuracy, balanced accuracy and the Kappa statistic from the fitted multinomial regression, RF and NN models on the validation set. Multinomial regression, with a higher accuracy of 0.630, slightly outperforms the NN model, which has an accuracy of 0.628. However, the NN model results in the highest Kappa statistic of 0.303 and the highest balanced accuracy of 0.5. All three models have a balanced accuracy of around 0.5 and a Kappa statistic of around 0.3, indicating room for improvement in predicting LMClass with imbalanced categories.

**Table 4:** Performance of multinomial regression, RF and NN models on the validation set for LMClass prediction.

| Model | Accuracy | Balanced accuracy | Kappa |
| --- | --- | --- | --- |
| Multinomial | 0.630 | 0.499 | 0.299 |
| RF | 0.619 | 0.494 | 0.290 |
| NN | 0.628 | 0.500 | 0.303 |

The information of selected predictors in the multinomial regression model for LMClass prediction is provided in Table A5 in Appendix A . In multinomial regression, we set the baseline class to be short stay & discharged, and the odds ratio with 95% CI for each selected predictor was collected for another two classes (e.g. short stay & deceased and long stay without specifying mortality outcomes) with respect to the baseline. The odds ratios of MODS for the class short stay & deceased and the class long stay without specifying mortality outcomes are 1.40 (95% CI = [1.34, 1.46]) and 1.15 (95% CI = [1.11, 1.19]), respectively. This means if one point increase in MODS score would increase the relative risk of short stay & discharged over short stay & deceased and long stay without specifying mortality outcomes by 40% (95% CI = [34%, 46%]) and 15% (95% CI = [11%, 19%]), respectively. Similarly, the odds ratios of NEMS for short stay & deceased and long stay without specifying mortality outcomes are 1.11 (95% CI = [1.10, 1.13]) and 1.07 (95% CI = [1.06, 1.08]), respectively. This means that one point increase in NEMS score would increase the relative risk of short stay & discharged over short stay & deceased and long stay without specifying mortality outcomes by 11% (95% CI = [10%, 13%]) and 7% (95% CI = [6%, 8%]), respectively, holding the other predictors fixed. As a reference, importance of predictors based on MDA is visualized in Figure B2 in the Appendix B which shows that both NEMS and MODS are important in LMClass prediction.

## 5 Conclusions and Discussion

In this work, we developed several models for health outcomes prediction in intensive care units. Compared with [16], adding the components of MODS and NEMS in the logistic regression for mortality prediction has an improvement of 3.5% in AUC values in the validation set (see Table 5). This study also demonstrates that MODS and NEMS with their components measured upon patient arrival significantly contribute to health outcome prediction in ICUs. In mortality prediction, achieving the highest sensitivity and NPV, RF outperforms logistic regression and NN, but logistic regression achieves the highest AUC. In LOS prediction, no big difference in the performance appears in the logistic regression and NN model. In practice, we need to evaluate the pros and cons of each model, and choose the best according to the type and goals of the analysis. For example, if we are concerned about correctly predicting the mortality outcomes among all the ICU patients, logistic regression is our first choice, while we would choose RF if we emphasize on prediction accuracy among the deceased patients. Explanation power may also play a role, especially with respect to the implications of such predictions. As an example, the predictors of long stays may help inform capacity planning and resource scheduling.

**Table 5:**
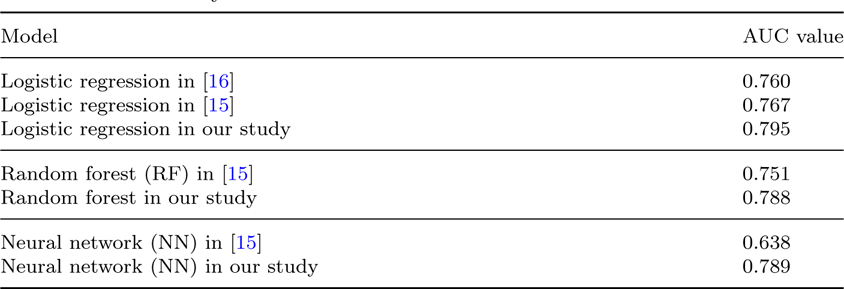
Comparison on AUC values in Mortality prediction between previous works and our study.

| Model | AUC value |
| --- | --- |
| Logistic regression in [16] | 0.760 |
| Logistic regression in [15] | 0.767 |
| Logistic regression in our study | 0.795 |
| Random forest (RF) in [15] | 0.751 |
| Random forest in our study | 0.788 |
| Neural network (NN) in [15] | 0.638 |
| Neural network (NN) in our study | 0.789 |

Furthermore, in terms of the definition of prolonged stay in the ICU, random forests and neural networks have greatly improved LOS prediction when we cut the short and long LOS at 5 days instead of 7 or 21 days as in [15]. A comparison on AUC values in LOS prediction between previous works and our study is provided in Table 6. However, as in [15] for LOS prediction, we find it is harder to classify a short or long stay than to detect mortality status. The underlying reason could be the definition of prolonged LOS as a binary health outcome. To improve the prediction accuracy, survival models can be developed for LOS prediction, and in this scenario LOS can be considered as a continuous time-to-event response.

**Table 6:**
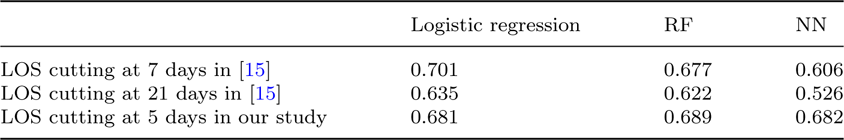
Comparison on AUC values in LOS prediction between work in [15] and our study

|  | Logistic regression | RF | NN |
| --- | --- | --- | --- |
| LOS cutting at 7 days in [15] | 0.701 | 0.677 | 0.606 |
| LOS cutting at 21 days in [15] | 0.635 | 0.622 | 0.526 |
| LOS cutting at 5 days in our study | 0.681 | 0.689 | 0.682 |

A trade-off between interpretation power and accuracy of prediction usually exists in predictive modelling. Logistic and multinomial regression models provide an interpretation for quantitative relationships between predictors and health outcomes using odds (i.e., relative risk). Compared with regression-based models, RF provides qualitative relationships using MDA, while NN is a black box whose statistical theoretical justifications are still under investigation in different frameworks [87, 88].

To our best of knowledge, we are the first to combine mortality with prolonged LOS to construct a new categorical health outcome and develop MODS and NEMS based predictive models for its prediction. In our expectation, more complexity occurs in this three-level outcome, making it more challenging to achieve high prediction accuracy. More complex deep learning models such as convolutional neural networks [89] and recurrent neural networks [90] can be applied but with higher computational costs.

It is important to note that, our data, with two main intensive care scoring systems, MODS and NEMS, were collected from two ICUs in London, Ontario, Canada, and the results may not be consistent with those in other ICUs outside of London, Ontario. For future work, a larger data set including the cases in several different ICUs from the Critical Care Information System in Ontario will be obtained for further analysis and validation based on [91]. COVID-19 patients will also be included in the new data set for predictive modelling.

## CRediT authorship contribution statement

**Chengqian Xian:** Conception and design of the study, Implementation of statistical analyses, Writing – original draft, Writing – review & editing. **Camila P.E. de Souza:** Conception and design of the study, Writing – review & editing. **Felipe F. Rodrigues** Conception and design of the study, Writing – review & editing.

## Data Availability

The authors do not have permission to share data.

## Acknowledgments

This research work is supported by the Natural Sciences and Engineering Research Council of Canada (NSERC). All authors approved the version of the manuscript to be published.

## Appendix A Tables

**Table A1:**
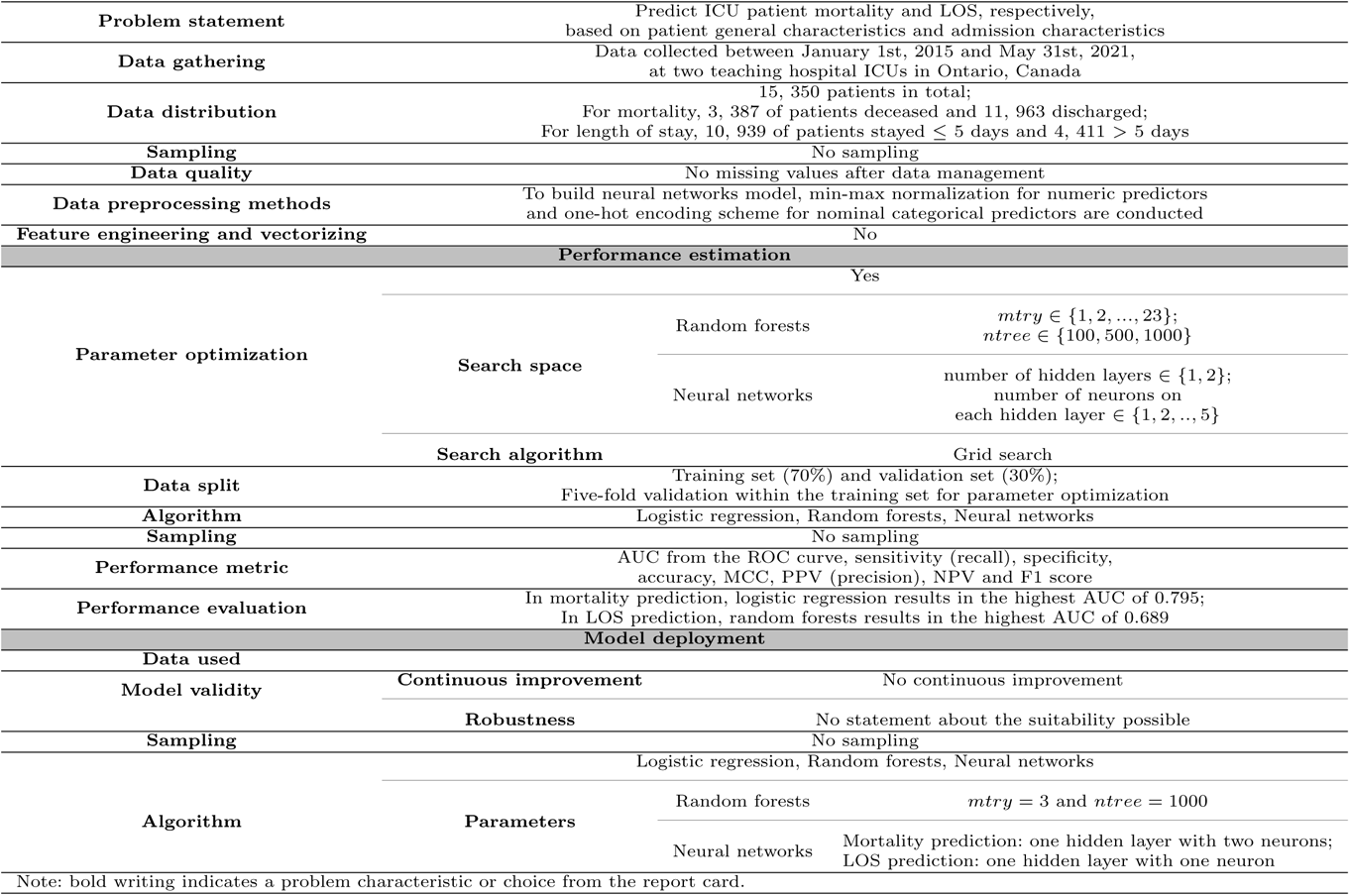
Report card based on the ICU data set for mortality and LOS predictions

**Table A2:**
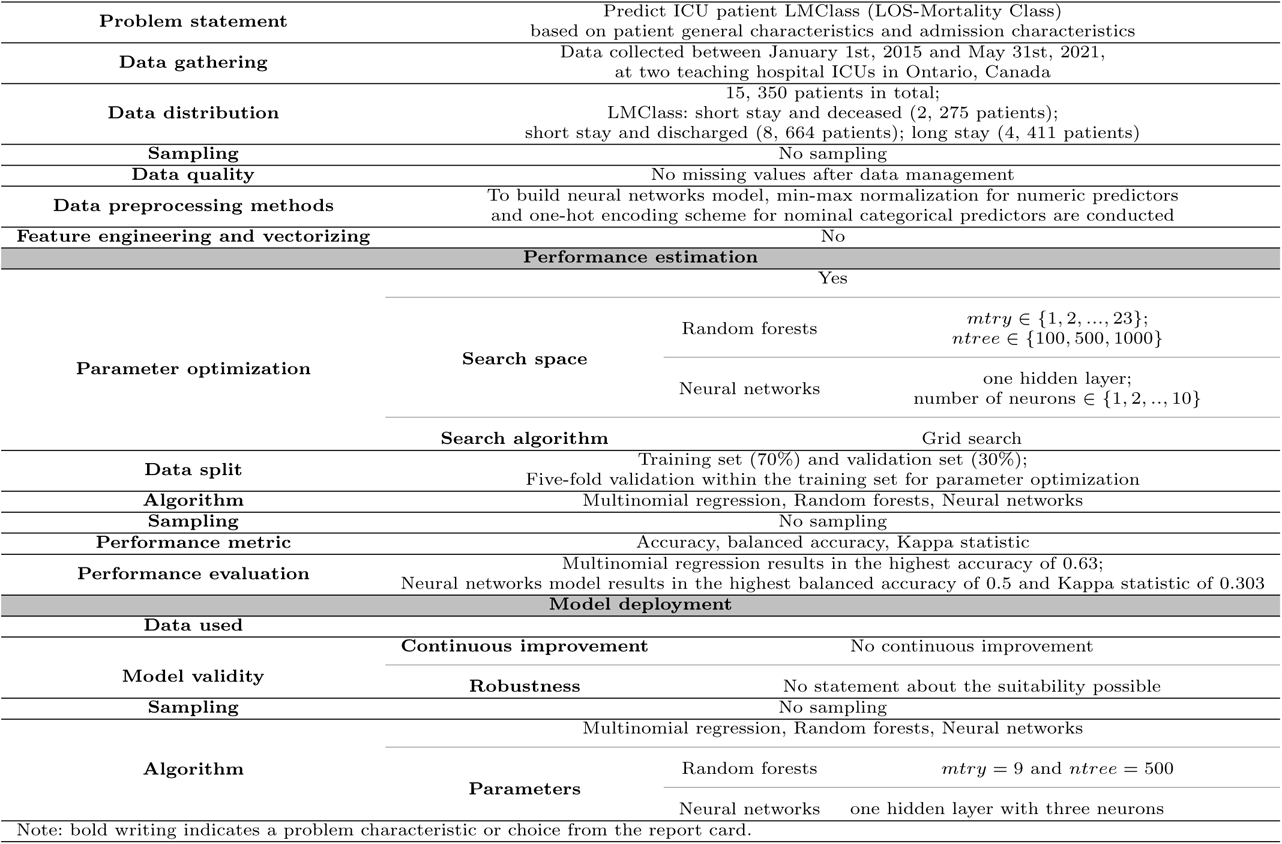
Report card based on the ICU data set for LMClass prediction

**Table A3:** Selected predictors in logistic regression for mortality prediction.

| Predictor | Coefficient | Odds ratio [95% CI] | LRT $p$-value<sup>1</sup> |
| --- | --- | --- | --- |
| Haematologic |  |  | < 0.0001 |
| None (reference) | - | - |  |
| Minimal | 0.38 | 1.46 [1.25, 1.71] |  |
| Mild | 0.45 | 1.57 [1.25, 1.96] |  |
| Moderate | 0.85 | 2.35 [1.82, 3.02] |  |
| Severe | 1.22 | 3.40 [2.22, 5.20] |  |
| Hepatic |  |  | < 0.0001 |
| None (reference) | - | - |  |
| Minimal | 0.13 | 1.14 [0.94, 1.39] |  |
| Mild | 0.27 | 1.30 [0.94, 1.80] |  |
| Moderate | 0.87 | 2.39 [1.57, 3.61] |  |
| Severe | 1.20 | 3.32 [2.14, 5.15] |  |
| Renal |  |  | < 0.0001 |
| None (reference) | - | - |  |
| Minimal | 0.30 | 1.36 [1.20, 1.53] |  |
| Mild | 0.49 | 1.64 [1.38, 1.95] |  |
| Moderate | 0.57 | 1.77 [1.37, 2.27] |  |
| Severe | 0.13 | 1.14 [0.86, 1.51] |  |
| Cardiovascular |  |  | < 0.0001 |
| None (reference) | - | - |  |
| Minimal | 0.27 | 1.31 [1.16, 1.47] |  |
| Mild | 0.57 | 1.77 [1.44, 2.17] |  |
| Moderate | 0.53 | 1.70 [1.24, 2.32] |  |
| Severe | 0.71 | 2.03 [1.22, 3.42] |  |
| Neurologic |  |  | < 0.0001 |
| None (reference) | - | - |  |
| Minimal | 0.28 | 1.32 [1.09, 1.61] |  |
| Mild | 0.13 | 1.14 [0.93, 1.40] |  |
| Moderate | 0.06 | 1.06 [0.84, 1.34] |  |
| Severe | 0.89 | 2.45 [2.13, 2.81] |  |
| Respiratory |  |  | < 0.0001 |
| None (reference) | - | - |  |
| Minimal | -0.07 | 0.94 [0.80, 1.09] |  |
| Mild | 0.13 | 1.14 [0.98, 1.33] |  |
| Moderate | 0.33 | 1.39 [1.20, 1.60] |  |
| Severe | 0.52 | 1.68 [1.37, 2.05] |  |

<sup>1</sup> $p$ -value of the likelihood ratio test for significance of the corresponding predictor

**Table A3 continued:** Selected predictors in logistic regression for mortality prediction.
| Predictor | Coefficient | Odds ratio [95% CI] | LRT $p$-value<sup>1</sup> |
| --- | --- | --- | --- |
| NEMS Score | 0.07 | 1.07 [1.06, 1.08] | < 0.0001 |
| Basic Monitoring |  |  | 0.005 |
| No (reference) | - |  |  |
| Yes | -2.23 | 0.11 [0.03, 0.48] |  |
| Intracranial Pressure Monitor |  |  | < 0.0001 |
| No (reference) | - | - |  |
| Yes | 0.74 | 2.09 [1.46, 2.95] |  |
| Dialysis |  |  | 0.002 |
| No (reference) | - | - |  |
| Yes | -0.44 | 0.65 [0.49, 0.85] |  |
| Intra-Aortic Balloon Pump |  |  | 0.119 |
| No (reference) | - | - |  |
| Yes | -0.40 | 0.67 [0.40, 1.11] |  |
| Other Interventions |  |  | 0.001 |
| No (reference) | - | - |  |
| Yes | -0.21 | 0.81 [0.72, 0.92] |  |
| Interventions Outside this Unit |  |  | < 0.0001 |
| No (reference) | - | - |  |
| Yes | -0.43 | 0.65 [0.57, 0.75] |  |
| Age | 0.03 | 1.03 [1.03, 1.04] | < 0.0001 |
| Sex |  |  | 0.001 |
| Female (reference) | - | - |  |
| Male | -0.17 | 0.84 [0.76, 0.94] |  |
| Patient Category |  |  | 0.001 |
| Medical (reference) | - | - |  |
| Surgical | -0.25 | 0.78 [0.67, 0.91] |  |
| Admission Source |  |  | < 0.0001 |
| Emergency (reference) | - | - |  |
| Operating Room | -0.94 | 0.39 [0.31, 0.48] |  |
| Outside Hospital/Other | -0.01 | 0.99 [0.86, 1.13] |  |
| Unit/Ward/Stepdown | 0.36 | 1.43 [1.23, 1.66] |  |
| Admission Diagnosis |  |  | < 0.0001 |
| Cardiovascular (reference) | - | - |  |
| Gastrointestinal | -0.45 | 0.64 [0.51, 0.80] |  |
| Neurological | -0.06 | 0.94 [0.79, 1.14] |  |
| Other | -0.58 | 0.56 [0.47, 0.66] |  |
| Respiratory | -0.48 | 0.62 [0.53, 0.72] |  |
| Trauma | -0.11 | 0.89 [0.68, 1.16] |  |

<sup>1</sup> $p$ -value of the likelihood ratio test for significance of the corresponding predictor

**Table A4:** Selected predictors in logistic regression for LOS prediction.

| Predictor | Coefficient | Odds ratio [95% CI] | LRT $p$-value<sup>1</sup> |
| --- | --- | --- | --- |
| MODS Score | 0.04 | 1.04 [1.02, 1.06] | 0.012 |
| Cardiovascular |  |  | 0.052 |
| None (reference) | - | - |  |
| Minimal | 0.09 | 1.09 [0.98, 1.21] |  |
| Mild | -0.19 | 0.83 [0.68, 1.00] |  |
| Moderate | -0.22 | 0.81 [0.59, 1.09] |  |
| Severe | -0.29 | 0.75 [0.45, 1.22] |  |
| Respiratory |  |  | < 0.0001 |
| None (reference) | - | - |  |
| Minimal | 0.12 | 1.12 [0.98, 1.28] |  |
| Mild | 0.29 | 1.34 [1.16, 1.53] |  |
| Moderate | 0.40 | 1.50 [1.30, 1.73] |  |
| Severe | 0.17 | 1.19 [0.97, 1.46] |  |
| NEMS Score | 0.04 | 1.04 [1.04, 1.05] | < 0.0001 |
| Central Venous Line |  |  | < 0.0001 |
| No (reference) | - | - |  |
| Yes | 0.46 | 1.59 [1.41, 1.79] |  |
| Arterial Line |  |  | 0.041 |
| No (reference) | - | - |  |
| Yes | 0.23 | 1.25 [1.12, 1.41] |  |
| Intracranial Pressure Monitor |  |  | 0.001 |
| No (reference) | - | - |  |
| Yes | 0.67 | 1.96 [1.43, 2.66] |  |
| Dialysis |  |  | 0.001 |
| No (reference) | - | - |  |
| Yes | -0.22 | 0.80 [0.64, 0.99] |  |
| Interventions Outside this Unit |  |  | 0.007 |
| No (reference) | - | - |  |
| Yes | -0.13 | 0.88 [0.78, 0.98] |  |

<sup>1</sup> $p$ -value of the likelihood ratio test for significance of the corresponding predictor

**Table A4 continued:** Selected predictors in logistic regression for LOS prediction.
| Predictor | Coefficient | Odds ratio [95% CI] | LRT $p$-value<sup>1</sup> |
| --- | --- | --- | --- |
| Sex |  |  | 0.026 |
| Female (reference) | - | - |  |
| Male | 0.11 | 1.12 [1.02, 1.23] |  |
| Admission Source |  |  | < 0.0001 |
| Emergency (reference) | - | - |  |
| Operating Room | -0.33 | 0.72 [0.62, 0.83] |  |
| Outside Hospital/Other | 0.47 | 1.61 [1.43, 1.81] |  |
| Unit/Ward/Stepdown | 0.34 | 1.40 [1.23, 1.60] |  |
| Admission Diagnosis |  |  | < 0.0001 |
| Cardiovascular (reference) | - | - |  |
| Gastrointestinal | 0.21 | 1.23 [1.01, 1.49] |  |
| Neurological | 0.23 | 1.25 [1.06, 1.49] |  |
| Other | 0.18 | 1.20 [1.03, 1.39] |  |
| Respiratory | 0.55 | 1.73 [1.51, 1.99] |  |
| Trauma | 0.86 | 2.37 [1.94, 2.90] |  |

<sup>1</sup> $p$ -value of the likelihood ratio test for significance of the corresponding predictor

**Table A5:**
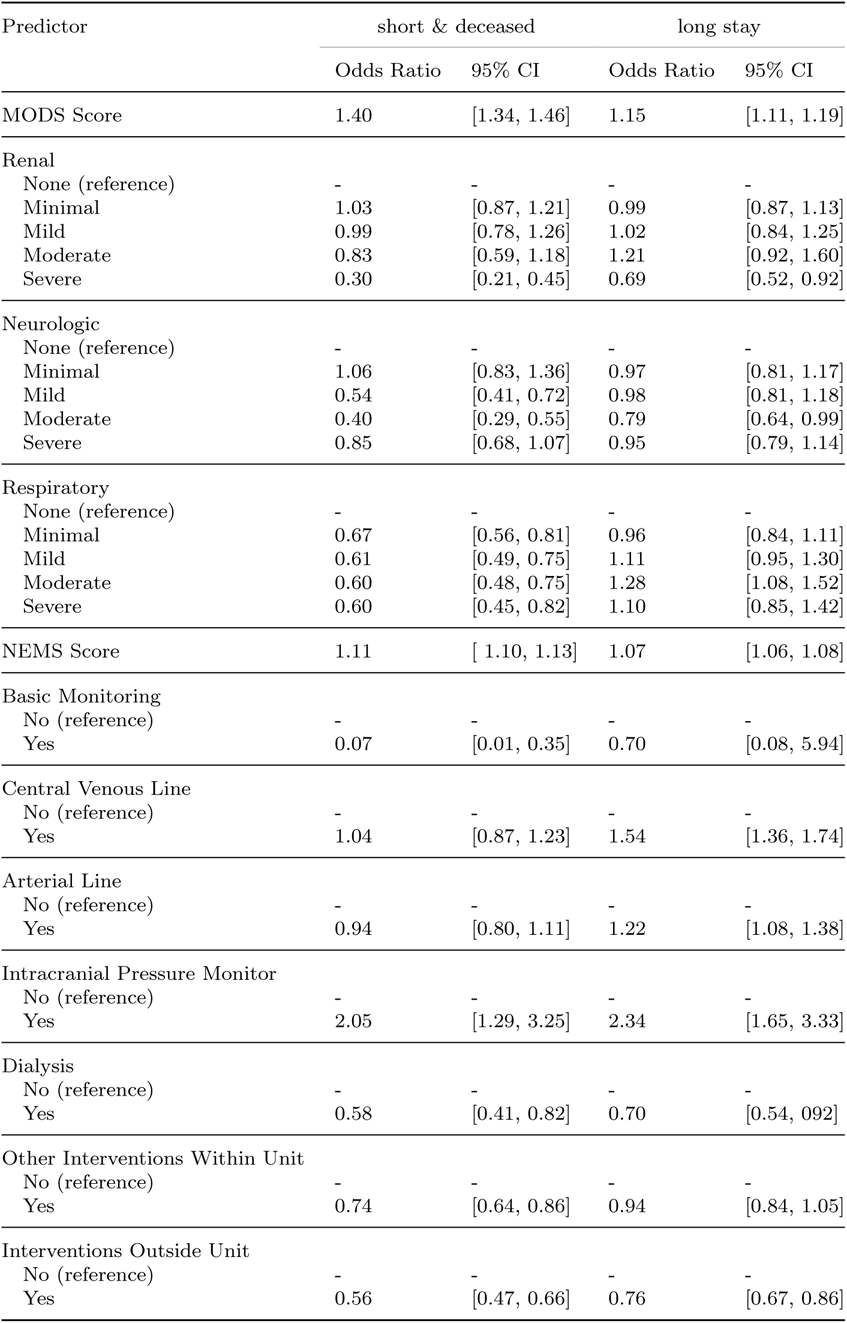
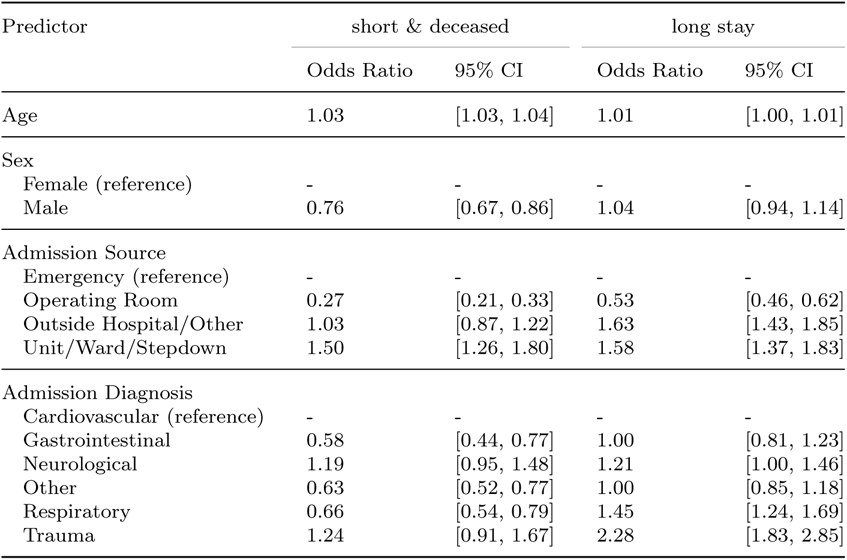
Selected predictors in multinomial regression for LMClass prediction.

**Table A6:** MODS Components (Adapted From Marshal et al 1995[10]).

| Organ System | Indicator of Dysfunction | Degree of Dysfunction |  |  |  |  |
| --- | --- | --- | --- | --- | --- | --- |
|  |  | None (0) | Minimal (1) | Mild (2) | Moderate (3) | Severe (4) |
| Respiratory | $PaO_2/FiO_2$ | $> 300$ | $226 - 300$ | $151 - 225$ | $76 - 150$ | $\leq 75$ |
| Renal | Creatinine ($mmol/L$) | $\leq 100$ | $101 - 200$ | $201 - 350$ | $351 - 500$ | $> 500$ |
| Cardiovascular | Pressure-adjusted rate | $\leq 10.0$ | $10.1 - 15.0$ | $15.1 - 20.0$ | $20.1 - 30.0$ | $> 30.0$ |
| Hematological | Platelets ($\times 10^3/mm^3$) | $> 120$ | $80 - 120$ | $50 - 80$ | $20 - 50$ | $< 20$ |
| Hepatic | Bilirubin ($\mu mol/L$) | $\leq 20$ | $21 - 60$ | $61 - 120$ | $121 - 240$ | $> 240$ |
| Neurological | Glasgow Coma Score | 15 | $13 - 14$ | $10 - 12$ | $7 - 9$ | $\leq 6$ |

**Table A7:** NEMS Components (Adapted From Miranda et al 1997 [12]).

| Item | Points |
| --- | --- |
| 1. *Basic monitoring*: hourly vital signs, regular record and calculation of fluid balance | 9 |
| 2. *Intravenous medication*: bolus or continuously, not including vasoactive drugs | 6 |
| 3. *Mechanical ventilatory support*: any form of mechanical/assisted ventilation, with or without PEEP (e. g., continuous positive airway pressure), with or without muscle relaxants | 12 |
| 4. *Supplementary ventilatory care*: breathing spontaneously through endotracheal tube; supplementary oxygen any method, except if (3) applies | 3 |
| 5. *Single vasoactive medication*: any vasoactive drug | 7 |
| 6. *Multiple vasoactive medication*: more than one vasoactive drug, regardless of type and dose | 12 |
| 7. *Dialysis techniques*: all | 6 |
| 8. *Specific interventions in the ICU*: such as endotracheal intubation, introduction of pacemaker, cardioversion, endoscopy, emergency operation in the past 24 h, gastric lavage; routine interventions such as X-rays, echocardiography, electrocardiography, dressings, introduction of venous or arterial lines, are not included | 5 |
| 9. *Specific interventions outside the ICU*: such as surgical intervention or diagnostic procedure; the intervention/procedure is related to the severity of illness of the patient and makes an extra demand upon manpower efforts in the ICU | 6 |
| Total | 56 |

## Appendix B Figures

**Fig. B1:**
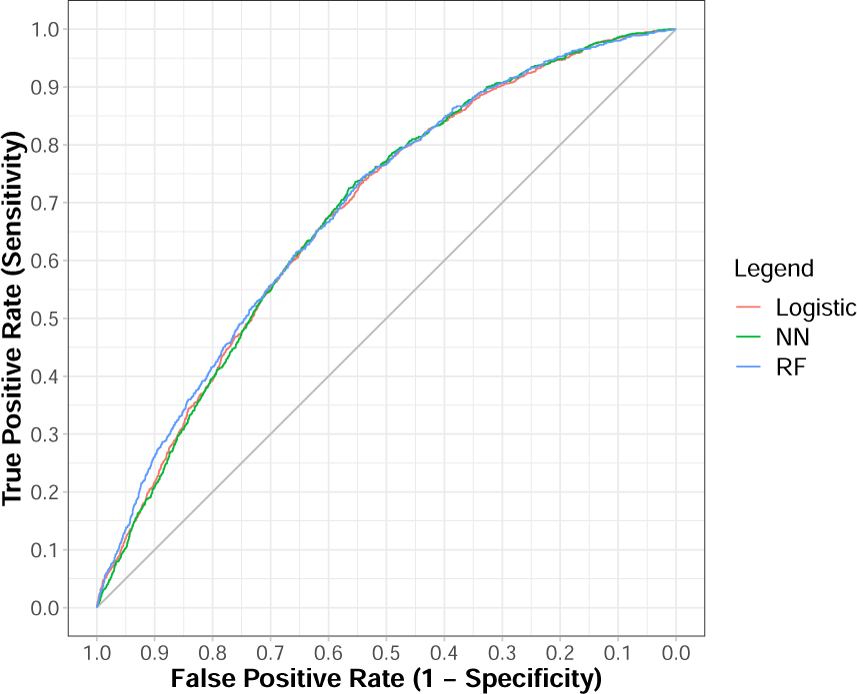
ROC curves for LOS prediction in validation set. Red: logistic regression; Green: random forests; Blue: neural networks

**Fig. B2:**
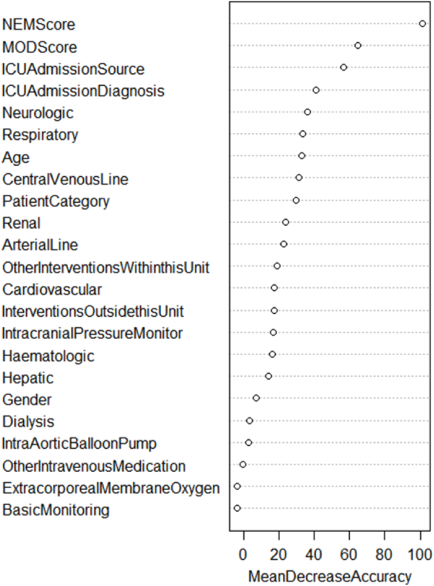
Importance of predictors based on RF model in LMClass prediction.

